# Decreased fentanyl potency as the primary driver of the 2024 decline in U.S. overdose deaths

**DOI:** 10.64898/2025.12.04.25341579

**Authors:** Daniel A. Busch

## Abstract

**Background:** In a profound reversal of prior trends, U.S. drug overdose deaths declined by 26.9% in 2024. Two proposed explanations are: (1) expansion of prevention, treatment, and harm-reduction infrastructure and (2) changes in the illicit fentanyl supply. This study evaluated which hypothesis best aligns with observed changes in drug involvement in overdose mortality.

**Methods:** CDC WONDER multiple-cause-of-death data for 2023 and 2024 were analyzed using complementary approaches. In a preliminary analysis, overdose deaths involving cocaine, methamphetamine, prescription opioids, heroin, and methadone were stratified by fentanyl involvement, and 2024/2023 mortality rate ratios were calculated. The primary analysis used a parsimonious 2×2 design (year × fentanyl involvement) to estimate differential mortality changes. A secondary analysis classified deaths into mutually exclusive strata defined by fentanyl, non-fentanyl opioid, and stimulant involvement, and estimated year-by-drug interaction effects using log-linear Poisson regression.

**Results:** Between 2023 and 2024, fentanyl-involved deaths declined by 36.5%; non-fentanyl-involved deaths declined by only 5.3% (p < 0.001). Regression models identified a large year × fentanyl interaction (RR = 0.65), consistent with a fentanyl-specific decline. In contrast, non-fentanyl opioid–involved (RR = 1.04) and stimulant-involved deaths (RR = 1.03) exhibited small relative increases.

**Conclusions:** The 2023–2024 decline in overdose mortality was confined to fentanyl-involved deaths. These findings are most consistent with supply-side changes affecting fentanyl toxicity rather than more uniform effects of infrastructure expansion. Continued investment in prevention and surveillance, with attention to potential market adaptation toward highly potent synthetic opioids, remains essential.

## 1. Introduction

A 27% decline in drug overdose deaths in the United States—from 110,037 in 2023 to 80,391 in 2024—constitutes a dramatic and historically unprecedented reversal of a two-decade upward trajectory (National Center for Health Statistics [NCHS], 2025). While this decrease is encouraging, its drivers and long-term stability remain uncertain. Fischer et al. (2025) proposed that recent mortality declines in Canada reflect contributions from four broad domains: changes in the at-risk population, drug supply characteristics, safer-use practices, and treatment access.

In the United States, two competing hypotheses have received particular attention. Supply-side explanations attribute the decline to reduced toxicity of the illicit drug supply—the “fentanyl hypothesis” (Drug Enforcement Administration [DEA], 2024; Milgram, 2024; Vangelov et al., 2026). This hypothesis draws support from DEA reports showing that declines in the purity of fentanyl in powder and counterfeit pills occurred at the same time as the decline in deaths (DEA, 2025).

In contrast, infrastructure-based explanations emphasize cumulative effects of expanded prevention, treatment, harm-reduction, and recovery services. Dasgupta et al. (2025) challenged the supply-side interpretation, citing state-level heterogeneity in overdose trends and methodological limitations in DEA purity estimates.

Not specifically raised by Dasgupta et al. (2025) is internal inconsistency in DEA reports. DEA (2024) reported the average purity of fentanyl powder as 19.2% in 2022 and 11.36% in 2024; however, these values are inconsistent with the purity trends shown in Figure 1. The reported figures correspond closely to purity levels in the final months of each year rather than to annual averages.

**Figure 1.**
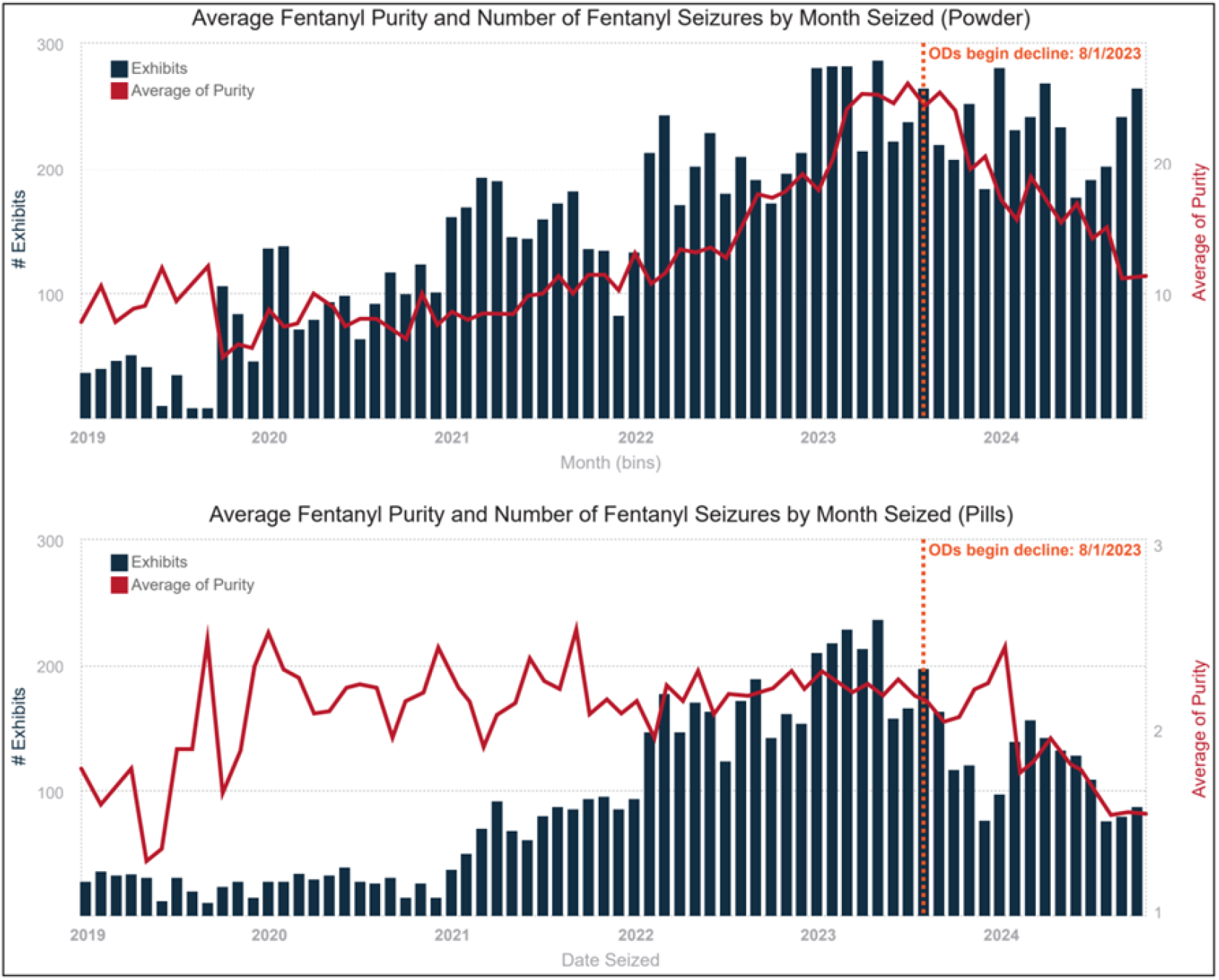
Fentanyl purity and seizures for pills and powder (DEA) Drug Enforcement Administration, 2024.

Centers for Disease Control and Prevention [CDC] analyses reported declines in overdose deaths across multiple drug categories, including fentanyl, cocaine, methamphetamine, prescription opioids, heroin, and methadone (CDC, 2025a; Anderer et al., 2025 (Figure 2).

**Figure 2.**
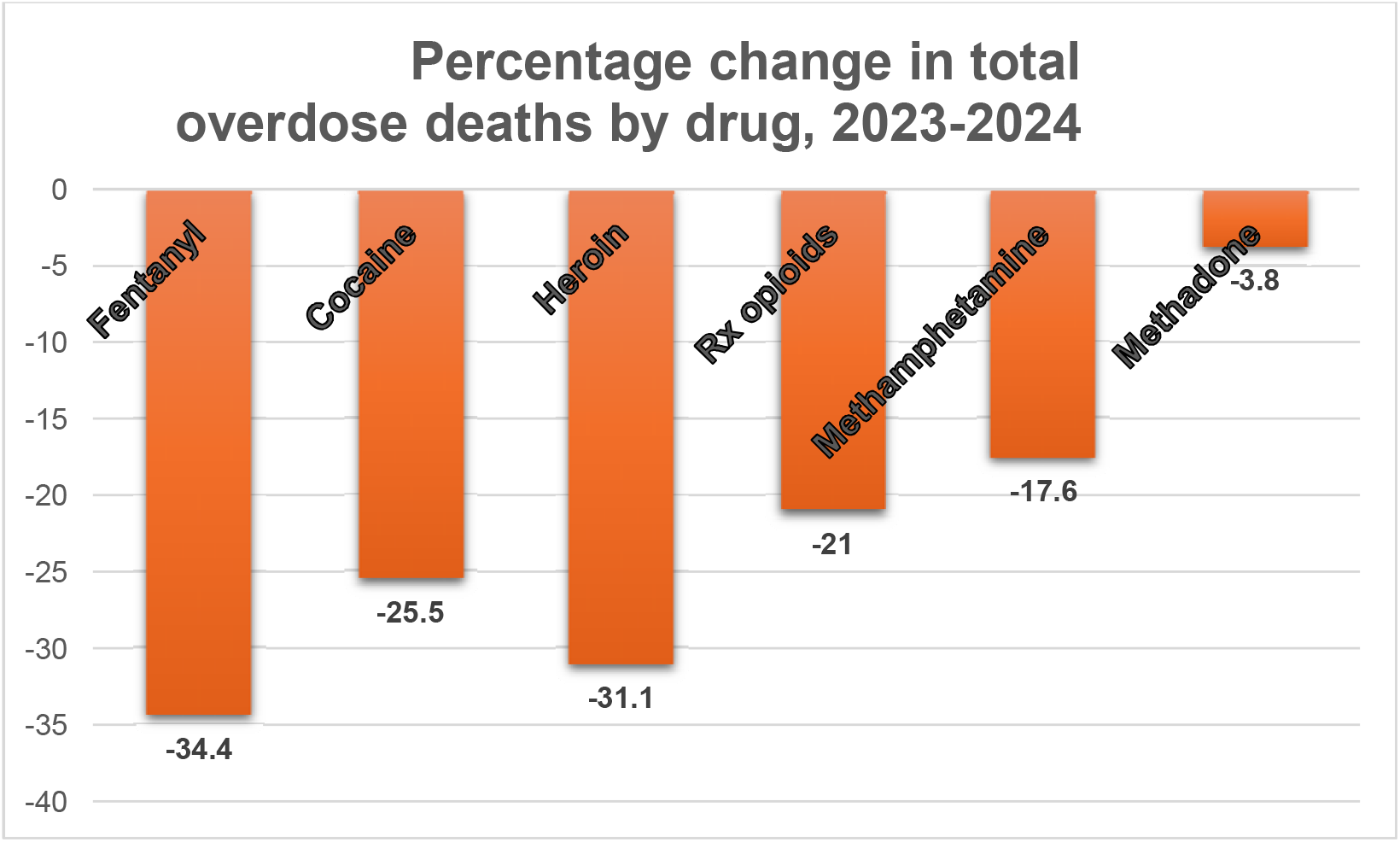
Percent change in total overdose deaths by drug, 2023-2024.

At face value, this broad decline appears inconsistent with the fentanyl hypothesis, which would predict reductions confined to fentanyl-involved deaths. However, the CDC’s methodology assigns each death to every drug involved in causing the death. As a result, a reduction in fentanyl toxicity could indirectly reduce deaths recorded for other drugs when fentanyl is co-involved, complicating the interpretation of drug-specific trends.

To disentangle these competing explanations, the present study examines drug-specific and drug-group-specific contributions to the 2023–2024 decline in overdose mortality. A preliminary analysis examines changes in deaths from 2023 to 2024 for five drugs stratified by fentanyl co-involvement. The main analysis examines 2023-2024 changes in fentanyl-involved deaths compared with non-fentanyl-involved deaths. A secondary analysis examines the relative contributions to the decline in mortality of three drug groups: (1) synthetic opioids other than methadone (primarily fentanyl), (2) non-fentanyl opioids (prescription opioids, methadone, and heroin), and (3) stimulants (primarily cocaine and methamphetamine).

This approach allows testing of specific hypotheses derived from the fentanyl and infrastructure hypotheses.

If reduction in fentanyl potency has been the primary driver of the mortality decline, decreases should be concentrated among fentanyl-involved deaths, with little or no decline in non-fentanyl-involved deaths.

If infrastructure expansion has been the primary driver, declines would be expected across drug classes. Since most infrastructure interventions—including naloxone distribution and medication for opioid use disorder (MOUD)—specifically target opioids, an infrastructure-driven decline should be more pronounced in opioid-involved deaths than in those involving stimulants. Effects of infrastructure expansion should be similar for fentanyl and non-fentanyl opioids, with the exception of fentanyl test strips, which would have a specific effect on fentanyl overdose deaths.

## 2. Methods

### 2.1. Data source

Mortality data were obtained from the CDC WONDER Multiple Cause of Death database. Deaths from 2018–2024 were identified using underlying cause-of-death codes for drug poisoning (ICD-10: X40–X44, X60–X64, X85, Y10–Y14). With the exception of the benzodiazepine analysis (Supplement 3), mortality data were downloaded in mid-2024. At that time, data for 2023 were not finalized, data for 2024 were provisional, and populations for 2023 and 2024 were the same.

### 2.2. Drug classification

Drug involvement was defined using ICD-10 multiple cause-of-death codes. “Fentanyl involvement” in the preliminary, primary, and secondary analyses was defined by code T40.4 (synthetic opioids other than methadone). “Non-fentanyl opioid involvement” was defined by T40.0–T40.3 (opium, heroin, other opioids (such as prescription opioids), and methadone. “Stimulant involvement” was defined by T40.5 (cocaine) and/or T43.6 (psychostimulants with abuse potential, including methamphetamine). Deaths coded as “other and unspecified narcotics” (ICD-10 T40.6) were excluded from primary fentanyl-involved and non-fentanyl-involved classifications, as this code reflects nonspecific opioid involvement and does not reliably indicate fentanyl absence (CDC, 2024). Polysubstance deaths were included and classified according to the presence or absence of each drug category; thus, a single death could contribute to multiple drug categories.

### 2.3. Study design and analytic framework

The primary objective was to determine whether the 2023–2024 decline in overdose mortality differed between deaths involving fentanyl and those not involving fentanyl. To address this question, a hierarchical analytic strategy was used, consisting of:

1. Preliminary descriptive analyses to examine fentanyl co-involvement within deaths involving five non-fentanyl drug categories (cocaine, methamphetamine, prescription opioids, heroin, and methadone).
2. Primary inferential analysis using a parsimonious 2 × 2 design (year x fentanyl involvement) to assess mortality shifts from 2023 to 2024.
3. Secondary multi-drug regression analyses incorporating stimulant and non-fentanyl opioid involvement as well as presence or absence of fentanyl involvement in overdose deaths in 2023 and 2024.
4. Sensitivity and supplementary analyses to evaluate robustness and explore alternative mechanisms.

This approach prioritized clarity of causal inference and minimized reliance on complex modeling assumptions.

#### 2.3.1 Preliminary analysis: fentanyl involvement within drug categories

Overdose deaths involving cocaine, methamphetamine, prescription opioids, heroin, and methadone were disaggregated into fentanyl-involved and non-fentanyl-involved subgroups. For each drug, 2024/2023 mortality rate ratios (RRs) were calculated and compared between fentanyl-involved and non-fentanyl-involved categories.

Aggregated analyses were conducted across all five drugs and across the three opioid drugs (prescription opioids, heroin, and methadone). Differences in log rate ratios were tested using standard z-tests.

Where needed, log-linear regression models were developed from 2018-2023 overdose death rates to predict 2024 deaths. For display purposes, logs of observed and predicted 2024 death rates were back-converted to numbers of deaths.

#### 2.3.2 Primary analysis: fentanyl versus non-fentanyl overdose deaths

The primary analysis used a parsimonious 2×2 design, defined by calendar year (2023 vs 2024) and fentanyl involvement (yes vs no). This design directly estimates the year × fentanyl interaction, which quantifies whether the magnitude of the mortality decline differed between fentanyl-involved and non-fentanyl-involved deaths. Within-group changes were summarized using 2024/2023 rate ratios (RRs). A log-linear Poisson regression model including main effects for year and fentanyl involvement and a prespecified year × fentanyl interaction was used to formally test whether the magnitude of the 2023–2024 change differed by fentanyl involvement. The interaction term represents a ratio of 2024/2023 rate ratios.

#### 2.3.3 Secondary analysis: Multi-drug regression

To examine whether the fentanyl-specific decline varied across polysubstance combinations, deaths were aggregated into 14 mutually exclusive cells based on calendar year (2023 vs. 2024) and binary indicators for three drug classes: fentanyl, non-fentanyl opioids, and stimulants. Log-linear Poisson regression models included main effects for year, fentanyl involvement, non-fentanyl opioid involvement, and stimulant involvement, along with prespecified year × drug and three-way interaction terms. The interaction terms tested whether year-over-year changes differed significantly across drug combinations after adjusting for co-involvement. Within-group rate ratios were used to quantify the magnitude of change for each specific drug category.

These models were intended to describe relative differences across drug categories, not to replace the primary fentanyl-involved vs. non-fentanyl-involved comparison.

#### 2.3.4 Sensitivity and supplementary analyses

First, a sensitivity analysis was performed to assess robustness to opioid misclassification. Deaths coded as T40.6 were reclassified as fentanyl-involved, and the primary and secondary models were re-estimated (Supplement 1).

Second, to distinguish acute changes from longer-term secular trends, log-linear regression models were fitted to annual overdose death counts from 2018–2023 for five drug categories, stratified by fentanyl involvement. These models were used to generate predicted 2024 deaths, which were compared with observed values (Supplement 2).

Third, to evaluate whether changes in benzodiazepine co-involvement contributed to the fentanyl-specific mortality decline, deaths were stratified by year, fentanyl involvement, and benzodiazepine involvement, yielding eight mutually exclusive cells. Log-linear Poisson regression models with population offsets were fitted, including prespecified Year × Fentanyl, Year × Benzodiazepine, and three-way interaction terms (Supplement 3).

## 3. Results

### 3.1 Preliminary analysis: fentanyl involvement and CDC drug categories

Disaggregating overdose deaths involving cocaine, methamphetamine, prescription opioids, heroin, and methadone by fentanyl involvement revealed a pattern markedly different from that observed in aggregated drug categories (as in Figure 2). Across cocaine, methamphetamine, prescription opioids, and methadone, deaths involving fentanyl declined sharply from 2023 to 2024, while deaths without fentanyl involvement remained stable or increased (Figure 3).

**Figure 3.**
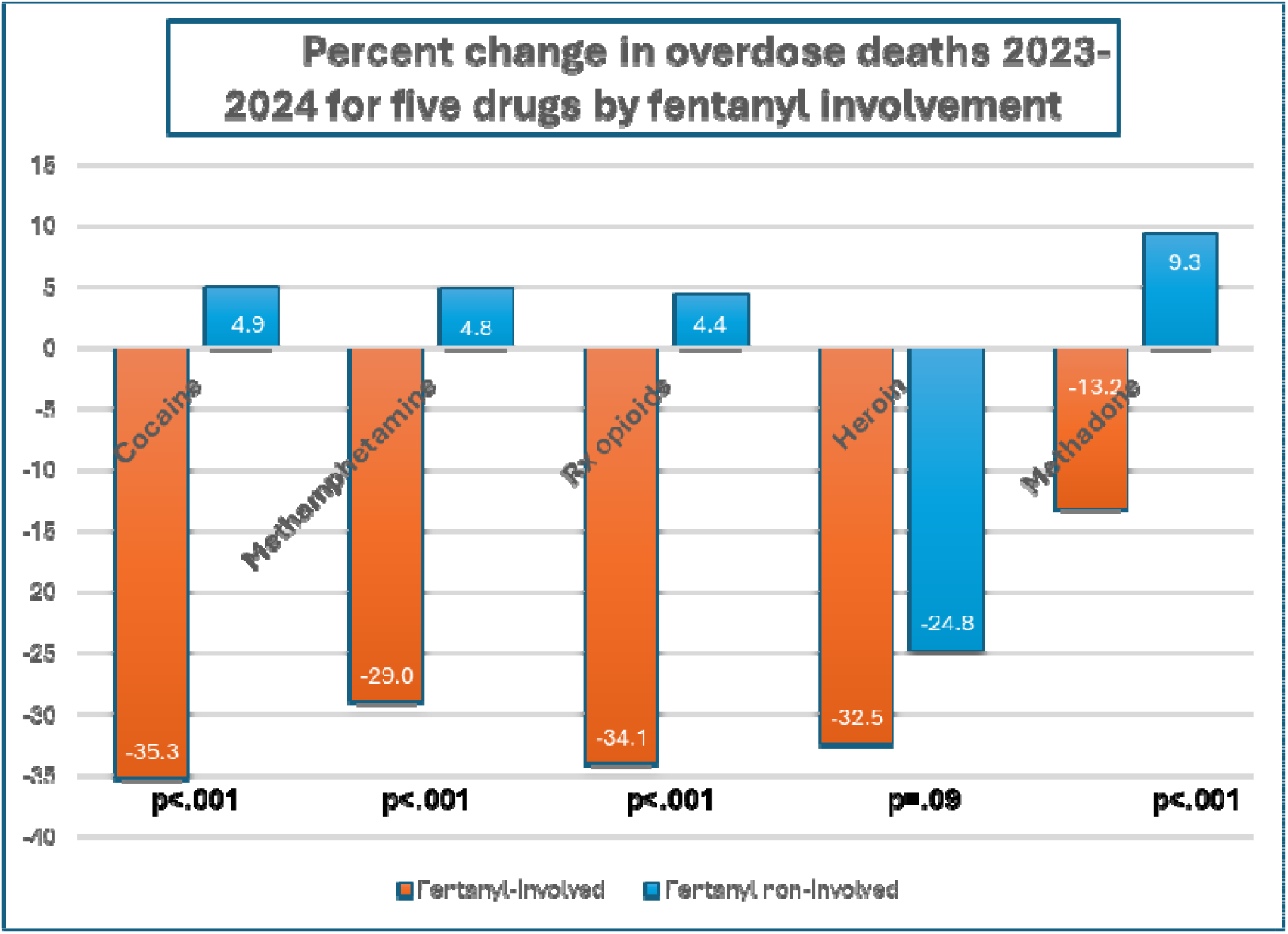

Differences in 2024/2023 mortality ratios for each fentanyl-involved and non-fentanyl-involved drug were highly significant (all p < 0.001). Heroin exhibited a similar but weaker pattern (p = 0.09).

When aggregated across all five non-fentanyl drug categories, fentanyl-involved deaths declined by 31.7%, whereas non-fentanyl-involved deaths increased by 2.6% (p < 0.05). When aggregated across the three non-fentanyl opioids examined here (prescription opioids, heroin, and methadone), fentanyl-involved deaths declined by 29.9%, while non-fentanyl-involved opioid deaths declined by 3.7% (p < 0.05). These findings indicate that the decreases reported by the CDC across multiple drug categories were driven almost entirely by reductions in fentanyl-involved deaths.

Because non-fentanyl-involved heroin deaths also declined from 2023 to 2024, supplementary trend-based analyses were conducted to distinguish acute changes from continuation of longer-term secular trends (Supplement 2). Projections based on 2018–2023 log-linear regression models showed that observed 2024 non-fentanyl-involved heroin deaths closely matched predicted values, while fentanyl-involved deaths across all drug categories fell far below projections. This pattern indicates that the 2023–2024 decline in non-fentanyl-involved heroin deaths reflected ongoing secular trends rather than the acute mortality decline observed for fentanyl.

### 3.2 Primary analysis: fentanyl versus non-fentanyl overdose deaths

Total overdose deaths declined from 110,037 in 2023 to 80,391 in 2024 (rate ratio [RR] = 0.729). This decline differed markedly by fentanyl involvement (Table 1).

**Table 1.** Overdose deaths in 2023 and 2024 by fentanyl-involvement.

| Table 1. Overdose deaths in 2023 and 2024 by fentanyl-involvement |  |  |  |
| --- | --- | --- | --- |
| Year | 2023 | 2024 | RR |
| Fentanyl-involved deaths | 76,282 | 48,422 | 0.635 |
| Non-fentanyl-involved deaths | 33,755 | 31,969 | 0.947 |
| Total overdose deaths | 110,037 | 80,391 | 0.731 |

In a parsimonious 2 × 2 analysis defined solely by calendar year and fentanyl involvement, fentanyl-involved overdose deaths declined by 36.5% from 2023 to 2024 (RR = 0.635), while deaths not involving fentanyl declined by 5.3% (RR = 0.947). This yielded a highly significant year × fentanyl interaction (ratio of RRs = 0.67, p < 0.001), indicating that the magnitude of mortality reduction was substantially greater for fentanyl-involved than for non-fentanyl-involved deaths.

This simple comparison directly addresses the primary study hypothesis and demonstrates that the national overdose mortality decline observed in 2024 was concentrated overwhelmingly among fentanyl-involved deaths.

### 3.3 Secondary analysis: multi-drug patterns and polysubstance involvement

To evaluate whether the decline in fentanyl-involved overdose deaths varied by combination with non-fentanyl opioids, stimulants, or their combination, overdose deaths were stratified by fentanyl, non-fentanyl opioid, and stimulant involvement and analyzed using log-linear Poisson regression models (Table 2).

**Table 2.**
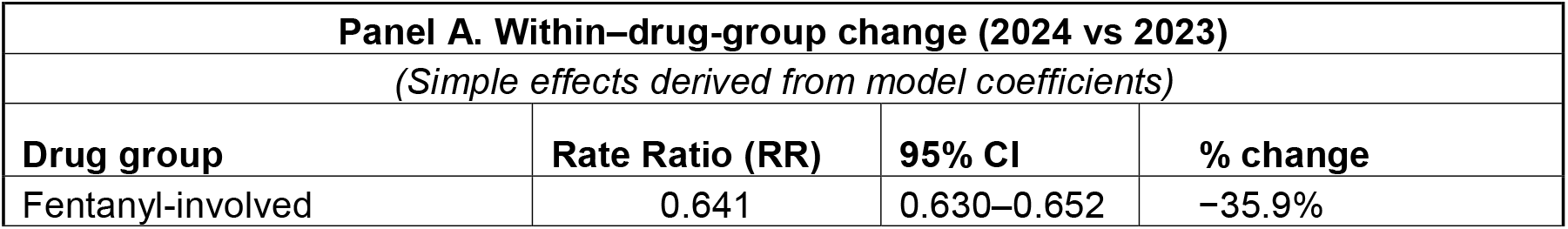
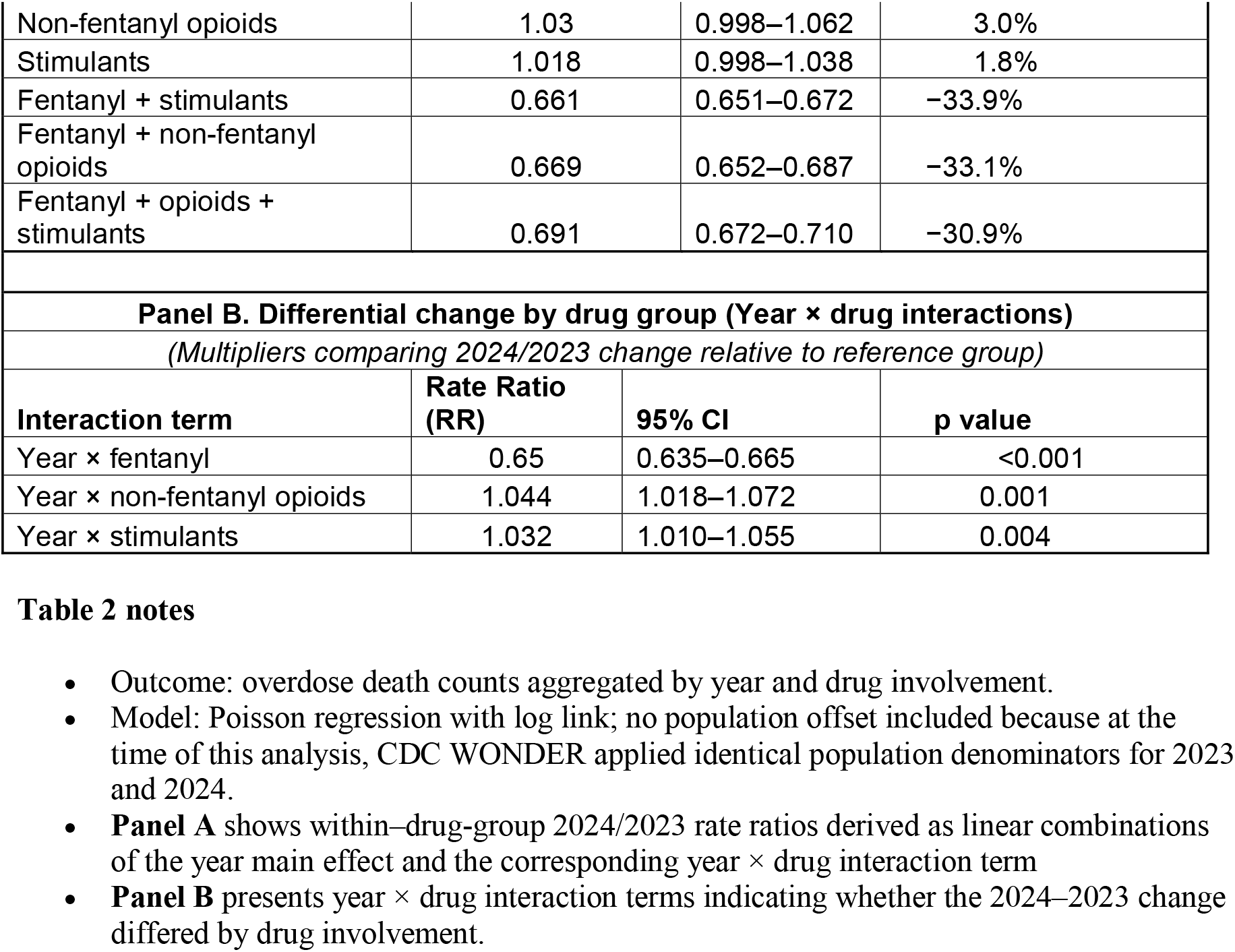
Changes in overdose deaths from 2023 to 2024 by drug involvement.

Across strata of fentanyl-involved overdose deaths, the 2024/2023 decline in fentanyl-involved overdose deaths was equivalent among all strata, including fentanyl alone (RR = 0.64), fentanyl combined with stimulants (RR = 0.66), fentanyl combined with non-fentanyl opioids (RR = 0.67), and fentanyl combined with both opioids and stimulants (RR = 0.69). In contrast, deaths not involving fentanyl showed little change, including small increases among non-fentanyl opioid–involved deaths (RR = 1.03) and stimulant-involved deaths (RR = 1.02).

In adjusted models, the year × fentanyl interaction remained dominant (RR = 0.65, p < 0.001), confirming that the mortality decline was specific to fentanyl involvement. In contrast, year × non-fentanyl opioid (RR = 1.04) and year × stimulant (RR = 1.03) interactions were minimally positive.

Together, these findings show that the 2023–2024 decline in overdose mortality was not shared equally across drug classes but was confined to fentanyl-involved deaths, irrespective of polysubstance involvement.

### 3.4 Sensitivity analyses and supplementary findings

Reclassification of deaths coded as “other and unspecified narcotics” (ICD-10 T40.6) as fentanyl-involved did not materially alter the direction, magnitude, or statistical significance of the primary findings (Supplement 1). Trend-based projections confirmed that the fentanyl-specific decline represented an acute deviation from secular trends, while non-fentanyl-involved deaths closely followed historical patterns (Supplement 2).

Analyses examining benzodiazepine co-involvement demonstrated that a reduction in benzodiazepine-involved deaths accounted for a small fraction of the decline in fentanyl-involved mortality (Supplement 3).

## 4.1. Discussion

This study was motivated by the question of whether the sharp decline in U.S. overdose deaths observed in 2024 was more consistent with supply-side changes in the illicit drug market or with the cumulative effects of investments in prevention, treatment, and harm-reduction infrastructure. To address this question without relying on DEA fentanyl purity data, CDC mortality data was used to conduct a population-level analysis of changes in overdose mortality by drug involvement.

Across all analyses, the 2023–2024 decline in overdose mortality was driven overwhelmingly by reductions in fentanyl-involved deaths. Because most prevention, treatment, and harm-reduction interventions do not distinguish between fentanyl and other opioids, an infrastructure-based explanation would be expected to produce comparable declines in fentanyl-involved and non-fentanyl opioid–involved deaths. This pattern was not observed. In secondary analyses separating fentanyl, non-fentanyl opioid, and stimulant involvement, the mortality decline was confined to fentanyl-involved deaths.

An initial descriptive analysis examined whether the decreases reported by the CDC for five major drug groups—methamphetamine, cocaine, prescription opioids, heroin, and methadone— differed by fentanyl involvement. For each drug individually, as well as for the aggregated group of five drugs and the subgroup of three opioids, the 2024/2023 mortality ratio was substantially lower when fentanyl was involved. In contrast, deaths without fentanyl involvement showed little change between 2023 and 2024.

The primary inferential analysis used a parsimonious 2×2 design defined by year and fentanyl involvement to directly test whether the 2023–2024 mortality decline differed between fentanyl-involved and non-fentanyl-involved overdose deaths. This approach provided a direct estimate of the year × fentanyl interaction with minimal modeling assumptions. At this ecological level, fentanyl-involved deaths declined by 36.5%, compared with a 5.3% decline in deaths not involving fentanyl.

Because this primary analysis did not address whether fentanyl-related declines varied across polysubstance combinations, a secondary regression analysis was conducted. This analysis classified overdose deaths into mutually exclusive categories defined by calendar year and the presence or absence of fentanyl, non-fentanyl opioids, and stimulants, allowing these effects to be estimated simultaneously while accounting for polysubstance involvement.

Results from this secondary analysis were fully consistent with findings from the primary analysis. The unprecedented decline in overdose mortality observed in 2024 was confined to fentanyl-involved deaths and was consistent across all polysubstance combinations containing fentanyl. In contrast, deaths involving non-fentanyl opioids, stimulants, or their combination in the absence of fentanyl showed minimal change.

Taken together, these findings provide strong evidence that supply-side changes affecting fentanyl toxicity—rather than rapid effects of infrastructure expansion—were the primary driver of the 2024 decline in overdose mortality. Although the specific mechanisms underlying these supply changes cannot be determined from these data, the results are compatible with DEA reports that documented declines in fentanyl purity in powder and counterfeit pills.

### 4.2. Alternative explanations

Alternative mechanisms could have contributed to the sharp 2023-2024 decline in overdose deaths.

#### 4.2.a. Alternative supply-side mechanisms

The DEA attributes declining overdose mortality to reduced fentanyl purity resulting from disruptions in precursor availability. While this interpretation is consistent with the observed decline in fentanyl-involved overdose mortality, inconsistencies in publicly reported DEA purity data necessitate consideration of additional supply-side explanations.

First, CDC WONDER classifies all synthetic opioids other than methadone under ICD-10 code T40.4. Decreased prevalence of highly potent synthetic opioids (e.g., carfentanil, nitazenes) in the illicit drug supply would result in decreased deaths coded T40.4 independent of changes in fentanyl purity.

Second, changes in the composition of sedative drugs co-occurring with fentanyl could modify overdose lethality. Decreased prevalence in the illicit drug supply of non-opioid depressants such as benzodiazepines, xylazine, and medetomidine would decrease the mortality rate with no change in fentanyl purity. Although xylazine and medetomidine lack unique codes, overdose deaths involving benzodiazepines are given a specific ICD-10 code (T42.4). A supplementary analysis (See Supplement 3) shows that the decline in the fentanyl-involved overdose mortality rate was considerably greater than the fall in the benzodiazepine-involved overdose mortality rate, indicating that a fall in the benzodiazepine mortality rate could not be the primary driver of the overall decline in overdose mortality from 2023 to 2024.

Third, a reduction in the proportion of illicit drug products containing fentanyl—rather than reduced fentanyl purity within individual doses—could theoretically reduce fentanyl-involved deaths while increasing deaths attributed to other drugs. However, this pattern was not observed:non-fentanyl-involved deaths declined by 5.3% rather than increasing, and there is no direct evidence that fentanyl prevalence among illicit drug products decreased during this period. Further evaluation of this hypothesis would require systematic drug market surveillance data, which are currently unavailable.

Finally, even if the immediate cause of decreased toxicity of the illicit drug supply is decreased fentanyl purity, the cause of the change in fentanyl purity cannot be determined from mortality data alone. While the DEA states that the reduction resulted from lack of precursor availability, other explanations—including deliberate reductions in fentanyl purity by suppliers for economic or strategic reasons—cannot be excluded.

#### 4.2.b Alternative Infrastructure-based explanations

Most infrastructure development measures, including MOUD and naloxone, should impact overdose deaths due to fentanyl and other opioids equally. In contrast, fentanyl test strips (FTS) could selectively reduce fentanyl-involved overdose deaths without similarly affecting deaths from other opioids. Bhai et al. (2025) reported that legalization of FTS was associated with an approximately 7% reduction in overall overdose mortality and a 13.5% reduction among Black individuals. However, other observational studies suggest more limited effects: Vickers-Smith et al. (2025) observed increased engagement in overdose risk-reduction behaviors among FTS users without detectable reductions in nonfatal overdose, and Kutcher et al. (2024) concluded that evidence for overdose prevention outcomes—particularly fatal overdose—remains limited relative to evidence for acceptability and behavioral change. Thus, while FTS may have contributed modestly to declining fentanyl-involved mortality, available evidence suggests they are unlikely to account for the magnitude of the observed decline.

### 4.3 Limitations

This study is limited by the ecological nature of the data, precluding assessment of local variation and individual-level mechanisms. The analysis lacks the granularity of DasGupta et al. (2025), which examined state-level differences in the timing of overdose declines and the effects of specific community-level interventions. Future research integrating state- and local-level changes in the drug supply with variation in prevention efforts, treatment availability, and harm reduction measures would provide a more complete understanding of the complex factors contributing to changes in overdose mortality.

At the time of all portions of this analysis except for Supplement 3, CDC mortality data for 2023 were not finalized, and mortality data for 2024 were provisional. However, CDC validation studies indicate that provisional counts reliably reflect final patterns (Ahmad et al., 2025). Death certificate data may underreport specific substances, but there is no evidence of differential misclassification between 2023 and 2024 sufficient to affect the study’s conclusions. Potential misclassification or underreporting of fentanyl involvement—particularly among deaths coded as T40.6—was addressed through sensitivity analyses, which yielded results in line with those of the primary study.

### 4.4. Policy Implications

Supply-side interventions have contributed to reduced fentanyl toxicity and warrant continued support. However, historical experience indicates that supply control has only limited long-term effectiveness, as illicit drug markets adapt through changes in precursor sourcing, production methods, trafficking routes, or substitution toward more potent synthetic opioids. Recent increases in deaths involving nitazenes (Bonner, 2024) and carfentanil (Tanz et al., 2024) underscore this risk.

Although this study does not support the hypothesis that infrastructure investments drove the rapid, fentanyl-specific mortality decline observed in 2024, such investments have contributed to longer-term reductions in deaths involving prescription opioids. Infrastructure development remains essential for sustained progress in addressing the opioid epidemic and should not be viewed as inconsistent with these findings.

Improved national surveillance of drug mortality and drug supply is needed. Routine disaggregation of overdose deaths by fentanyl co-involvement would enhance attribution of emerging trends, and expanded monitoring of high-potency synthetic opioids—including nitazenes and carfentanil—is critical given the potential for rapid market substitution.

Regular, accurate, and granular release of DEA data would be a major contribution to public health efforts.

## Supporting information

3.4 Sensitivity analyses and supplementary findings

3.4 Sensitivity analyses and supplementary findings

3.4 Sensitivity analyses and supplementary findings

## Data Availability

All data produced in the present study are available upon reasonable request to the authors

