## Supplementary material for "Decreased fentanyl potency as the primary driver of the 2024 decline in U.S. overdose deaths": 3.4 Sensitivity analyses and supplementary findings

**Supplemental File 1. Sensitivity analysis: Reclassification of T40.6 deaths as fentanyl-involved**

**Rationale**

In the primary analysis, deaths coded as “other and unspecified narcotics” (ICD-10 T40.6) were excluded from both fentanyl-involved and non-fentanyl-involved categories because this code does not reliably indicate fentanyl exposure. Because advances in postmortem toxicology suggest that many T40.6 deaths involve fentanyl, a sensitivity analysis was conducted to evaluate the robustness of findings to potential undercounting of fentanyl involvement.

**Methods**

For the sensitivity analysis, fentanyl involvement was redefined as the presence of ICD-10 T40.4 (synthetic opioids other than methadone) and/or T40.6. All other analytic procedures, including model specification and hypothesis testing, were identical to those used in the primary analysis.

**Results**

Reclassification of T40.6 deaths did not materially alter the direction, magnitude, or statistical significance of any primary findings (Table S1). Fentanyl-involved deaths continued to exhibit large declines from 2023 to 2024 (RR = 0.642; 95% CI: 0.631–0.653; p < 0.001), with a strong Year × Fentanyl interaction (RR = 0.650; 95% CI: 0.635–0.665; p < 0.001). Non-fentanyl-involved deaths showed minimal change. These findings indicate that undercounting of fentanyl involvement due to T40.6 coding does not account for the observed fentanyl-specific decline in overdose mortality.

| Table S1: Comparison of 2024/2023 mortality ratios (RR)for primary analysis | | |
| --- | --- | --- |
| (fentanyl = T40.4 only) and sensitivity analyses (fentanyl = T40.4 and/or T40.6) | | |
| *Results from log-linear Poisson regression models* | | |
| Quantity | Primary (T40.4 only) | Sensitivity (T40.4 ∪ T40.6) |
| Year RR (no fentanyl) | 1.03 | 1.03 |
| Fentanyl main effect | 1.97 | 1.98 |
| Year × fentanyl RR | 0.64 | 0.64 |
| Fentanyl-specific decline | 34.10% | 34.00% |
