## Supplementary material for "Decreased fentanyl potency as the primary driver of the 2024 decline in U.S. overdose deaths": 3.4 Sensitivity analyses and supplementary findings

**Supplemental File 2. Trend-based projections of fentanyl-involved and non-fentanyl-involved overdose deaths**

**Rationale**

The preliminary analysis demonstrated that, with the exception of heroin, non-fentanyl-involved overdose deaths among five drugs did not decline from 2023 to 2024. Because non-fentanyl-involved heroin mortality had exhibited a sustained decline from 2018–2023, a supplementary analysis was conducted to distinguish acute 2023–2024 changes from continuation of longer-term secular trends.

**Methods**

Log-linear regression models were fitted to annual overdose death counts from 2018–2023 for cocaine, methamphetamine, prescription opioids, heroin, and methadone, stratified by fentanyl involvement. These models were used to generate predicted 2024 death counts. One-sample, one-tailed t-tests assessed whether observed 2024 mortality differed from predicted values. Paired t-tests compared deviations between fentanyl-involved and non-fentanyl-involved groups. For display, predicted log values were back-transformed to counts.

**Results**

Observed 2024 fentanyl-involved overdose deaths fell markedly below predicted values for cocaine, methamphetamine, and prescription opioids (36–51% of predicted; all p < 0.05), with fentanyl-involved heroin and methadone deaths trending similarly. In contrast, observed non-fentanyl-involved deaths for all drugs closely matched predictions (89–115% of predicted; all p = NS) (Table S2).

| Table S2. Actual 2024 overdose deaths for the individual fentanyl-involved and non-fentanyl-involved drugs compared with deaths predicted by 2018-2023 log-linear regression equations | | | | | | |
| --- | --- | --- | --- | --- | --- | --- |
|  |  |  |  | Lower Prediction |  | Actual as % of |
|  |  | Actual | Predicted | Limit | p<.05 | predicted |
| 4. A. Fentanyl-only | | 16,072 | 39,081 | 21,125 | * | 41.1 |
| 4. B. Fentanyl-involved drugs | |  |  |  |  |  |
| Cocaine |  | 14,396 | 29,955 | 23,343 | * | 48.1 |
| Methamphetamine | | 16,366 | 45,202 | 21,143 | * | 36.2 |
| Prescription opioids | | 3,742 | 7,402 | 3,993 | * | 50.6 |
| Heroin |  | 2,208 | 3,120 | 1,830 |  | 70.8 |
| Methadone | | 1,694 | 2,605 | 1,654 |  | 65.0 |
| 4. C. Non-fentanyl-involved drugs | |  |  |  |  |  |
| Cocaine |  | 7,548 | 7,243 | 6,036 |  | 104.2 |
| Methamphetamine | | 12,354 | 13,898 | 10,096 |  | 88.9 |
| Prescription opioids | | 4,242 | 3,997 | 3,781 |  | 106.1 |
| Heroin |  | 536 | 530 | 272 |  | 101.1 |
| Methadone | | 1,533 | 1,336 | 1,062 |  | 114.7 |

**Table S2** Actual versus predicted 2024 overdose deaths for fentanyl-involved and non-fentanyl-involved drugs.

As a group, fentanyl-involved drug deaths were significantly lower than predicted (t = −5.40, df = 4, p < 0.005), whereas non-fentanyl-involved deaths were not (t = 0.63, df = 4, p = NS). The paired difference between fentanyl-involved and non-fentanyl-involved groups was highly significant (t = −7.11, df = 4, p < 0.001).

For heroin, observed 2024 non-fentanyl-involved deaths closely matched projected values, indicating that the 2023–2024 decline in non-fentanyl-involved heroin mortality reflects continuation of a pre-existing secular trend rather than the fentanyl-specific mortality shock.

**Interpretation**

These findings reinforce the conclusion that the sharp 2023–2024 decline in overdose mortality was specific to fentanyl-involved deaths and not shared by non-fentanyl-involved drugs.
