## Supplementary material for "Decreased fentanyl potency as the primary driver of the 2024 decline in U.S. overdose deaths": 3.4 Sensitivity analyses and supplementary findings

**Supplemental File 3. Impact of benzodiazepine involvement on fentanyl-related overdose mortality**

**Rationale**

Because benzodiazepines potentiate opioid-induced respiratory depression, a sharp reduction in benzodiazepine co-involvement could contribute to declines in fentanyl-involved overdose deaths. This supplementary analysis examined whether changes in benzodiazepine involvement could plausibly account for the magnitude of the fentanyl-specific mortality decline.

**Methods**

CDC WONDER data downloaded January 24, 2026, were analyzed. Deaths were stratified by year (2023 vs 2024), fentanyl involvement (present vs absent), and benzodiazepine involvement (present vs absent), yielding eight mutually exclusive strata.

Log-linear Poisson regression models with population offsets (2023: 334,914,895; 2024: 340,110,988) estimated main effects and interaction terms for Year, Fentanyl, Benzodiazepines, Year × Fentanyl, Year × Benzodiazepines, and the three-way interaction (Year × Fentanyl × Benzodiazepines). Rate ratios (RRs) with 95% confidence intervals (CIs) are reported.

**Results**

From 2023 to 2024, non-fentanyl-involved benzodiazepine deaths declined by 6.2%. Because 89.8% of fentanyl-involved deaths in 2023 did not involve benzodiazepines, even a comparable reduction among fentanyl-involved benzodiazepine deaths would have produced only a minimal effect on overall fentanyl mortality.

Within-stratum analyses showed that fentanyl-involved deaths declined more sharply when benzodiazepines were absent (RR = 0.648; 95% CI: 0.639–0.657) than when present (RR = 0.751; 95% CI: 0.725–0.778). Deaths not involving fentanyl showed minimal change regardless of benzodiazepine involvement (Table S3).

Regression analyses confirmed a dominant Year × Fentanyl interaction (RR = 0.656; 95% CI: 0.646–0.667; p < 0.001), a modest independent Year × Benzodiazepine effect (RR = 0.949; 95% CI: 0.917–0.982; p = 0.002), and a strong positive three-way interaction (RR = 1.226; 95% CI: 1.171–1.283; p < 0.001), indicating that benzodiazepine co-involvement attenuated the magnitude of the fentanyl-specific mortality decline.

**Interpretation**

Reductions in benzodiazepine involvement accounted for only a small fraction of the overall decline in fentanyl-involved overdose deaths. Moreover, fentanyl deaths involving benzodiazepines declined less than those without benzodiazepines, suggesting that sedative co-exposure blunted the mortality improvement associated with declining fentanyl toxicity.

|  | Table S3. Changes in overdose mortality by fentanyl and benzodiazepine involvement, 2023–2024 | | | | | | | |
| --- | --- | --- | --- | --- | --- | --- | --- | --- |
|  | *(Simple effects: 2025/2024 rate ratios calculated within each fentanyl–benzodiazepine stratum)* | | | | | | | |
|  | Results from log-linear Poisson regression with population offsets | | | | | | | |
|  | Panel A. Within–drug-group changes (2024 vs 2023) | | | | | | |  |
|  | Fentanyl | Benzodiazepines | | Rate Ratio (RR) | | 95% CI | | % Change |
|  | **+** | **+** | | 0.751 | | 0.725–0.778 | | −24.9% |
|  | **+** | **−** | | 0.648 | | 0.639–0.657 | | −35.2% |
|  | **−** | **+** | | 0.938 | | 0.900–0.979 | | −6.2% |
|  | **−** | **−** | | 0.988 | | 0.974–1.002 | | −1.2% |
| Panel B. Differential change by drug involvement (interaction effects) | | | | | | | | |
| Interaction term | | | Rate Ratio (RR) | | 95% CI | | p value | |
| Year × fentanyl | | | 0.656 | | 0.646–0.667 | | <0.001 | |
| Year × benzodiazepines | | | 0.949 | | 0.917–0.982 | | 0.002 | |
| Year × fentanyl × benzodiazepines | | | 1.226 | | 1.171–1.283 | | <0.001 | |

Table Notes

- Outcome: overdose death rates.
- Model: Poisson regression with log link and population offsets for 2023 (334,914,895) and 2024 (340,110,988).
- Panel A shows within-stratum 2024/2023 mortality rate ratios.
- Panel B shows multiplicative interaction terms testing whether year-to-year mortality changes differed by fentanyl and benzodiazepine involvement.
